# Reference Values for Epicardial Adipose Tissue: Data from 27,500 CT scans in the General Population and Symptomatic Patients

**DOI:** 10.64898/2026.04.24.26351713

**Authors:** David Molnar, Chunliang Wang, Teemu Maaniitty, Elias Björnson, Martin Adiels, Carl-Johan Carlhäll, Tomas Jernberg, Joel Kullberg, Ellen Ostenfeld, Stefan Söderberg, Antti Saraste, Juhani Knuuti, Göran Bergström

**Author notes:** these authors contributed equally (David Molnar, Chunliang Wang). **Conflicts of interest statement: there’s no financial/personal interest or belief of any of the authors that would affect their objectivity.**. corresponding author: David Molnar, Turku PET Centre, Turku University Hospital, University of Turku, Kiinamyllynkatu 4-8, 20520 Turku, Finland.

## Abstract

**Background:** Increased epicardial adipose tissue volume (EATV) is a potentially important risk marker for coronary artery disease (CAD) available from cardiac computed tomography (CT) images. Sex-differences and effects of age and body size on EATV have been insufficiently explored, and no reliable reference values exist. Consequently, EATV has yet to find its deserved use in clinical practice.

**Objectives:** To define normal values by sex and age, the best normalization procedure for EATV to neutralize effects of body-size, explore the relationship between normalized EATV and cardiac risk, and propose a clinically meaningful cut-off.

**Methods:** AI-based automated EATV data from the general population (n=25,155) and a clinical cohort (n=2,482) with suspected CAD was normalized to height, BSA and heart volumes. Correlation between EATV and EAT attenuation was tested with Spearman’s rank correlation and linear regression to find the optimal normalization. Normalized EATV was compared to high-risk by SCORE2 and obstructive CAD in the population cohort. A cut-off including 95% of cases with obstructive CAD was defined in the general population and tested in the clinical cohort.

**Results:** EATV varied with sex and age across cohorts. Normalization of EATV to total heart volume (EATVh) was superior by all metrics and neutralized the effects of sex. High-risk by SCORE2 and the prevalence of obstructive CAD increased over quartiles of EATVh in the population cohort, and significantly higher EATVh was seen with obstructive CAD in both cohorts. A cut-off of 0.1 in EATVh had a negative predictive value for obstructive CAD of 97.1% in the general population and 88.9% in the clinical cohort.

**Conclusions:** EATV varies considerably with sex, age and body size. Normalization to heart volume outperformed other procedures, and EATVh is a useful marker of obstructive CAD in both the general population and symptomatic patients.

## Introduction

The epicardial adipose tissue (EAT) has been increasingly recognized as a predictor of coronary artery disease (CAD) including myocardial infarction^1–3^. The volume of EAT (EATV) and its attenuation (EATA), or radiodensity, can be reliably quantified from computed tomography (CT) images, both non-contrast enhanced (NCCT) and contrast enhanced angiographic images (CTA) ^4–7^.

Initially, definition of the EAT was performed manually, but recent advances in artificial intelligence based image processing has yielded numerous models which are capable of both semi-automatic and fully automatic delimitation of the EAT with precision comparable to manual expert work^8–10^.

However, despite intense research in the field, normal values have yet to be defined for the EAT, both for its volumes and attenuation values. The potential for establishing normal values has hitherto been reduced by most studies being small, selective of symptomatic patients, and highly variable technical performance of EAT measurements **[Supplement: Table S.1]**. Consequently, there is a conspicuous scarcity of reported data on sex differences in EATV and EATA^11,12^, and also a general paucity of data on the effects of age on EAT, despite age being previously shown to increase EATV in serial measurements over time^13,14^.

Notably, there is substantial co-linearity between EATV and adiposity and EATV and body size^15^. Pathophysiologically, a relative expansion of the EAT, which is seen in obesity^16–19^, is of interest as a risk marker, putatively driven by increased storage of lipids in the adipocytes. An increase in relative lipid content of the EAT will invariably result in a lower radiodensity, and a lower (more negative) mean EATA. While heart size correlates strongly with sex and general body size, the mean EATA should remain influenced mainly by the volumetric expansion in adipocytes. In contrast, a tall and lean person with a thin, normal layer of EAT but around a bigger heart could have the same total EATV as a short, but obese person with a thicker layer of EAT around a smaller heart. Consequently, to increase the specificity of EATV for true volumetric expansion, EATV would need to be normalized to a more reliable marker such as anatomical heart size.

The aims of the present study were: a) to define normal reference values for EATV and EATA in an unselected general population of ∼25,000 individuals (50% women), b) to explore the best model for normalizing EATV to body size, and, c) to compare EAT data from ∼2,500 symptomatic patients with suspected CAD to these reference values.

## Methods

### Informed consent and ethical permits

The study in its entirety conforms to the Declaration of Helsinki.

The unselected general population study, the “SCAPIS”, was approved (# 2010-228-31M) as a multi-center study by the ethical review board in Umeå, c/o Department of Medical Research, Umeå University, 901 87 Sweden. The current sub-study was approved by the Ethics Review Authority, Uppsala, Department 2 Medicine, Dnr 2024-02853-01. All participants have given written informed consent.

The study of clinical patients with suspected CAD was approved by the Ethics Committee of the Hospital District of Southwest Finland with no requirement for written informed consent due to the observational study design.

### Study populations

The general population cohort (“population cohort”) is a subset of the SCAPIS study, consisting of 25,155 participants with available CTA and EAT data (20 cases of incomplete images excluded) as well as complete clinical background data. In short, SCAPIS is a general-population-based prospective study (www.scapis.org), to which 30,154 men and women aged 50 to 64 years were randomly recruited from the census registry at six Swedish university sites (Gothenburg, Linköping, Malmö/Lund, Stockholm, Umeå, and Uppsala) between 2013 and 2018. The details of SCAPIS have been extensively described elsewhere^20^.

The clinical patient study cohort (“clinical cohort”) is a subset of a cohort of symptomatic patients evaluated with coronary CTA for suspected CAD between 2007 and 2017 at the Turku University Hospital in Finland (n=3,269) and comprises 2,482 patients with available data on EAT and heart volumes. Patients with previously known obstructive CAD or prior myocardial revascularization were not included in the original study cohort. Demographic and clinical data was obtained from electronic medical records.

### Computed tomography imaging Population cohort

Imaging was performed according to the SCAPIS protocol, using the same CT-scanners and protocols, Siemens Somatom Definition Flash with a Stellar detector (Siemens Healthcare, Forchheim, Germany). Care Dose 4D was used for dose optimization. NCCT image acquisition was ECG-gated, with tube voltage of 120 kV, and refmAs of 80 for calcium scoring. A matrix of 512×512 voxels in the axial plane was used, with a square DFOV in the range of 170-200 mm. Images were reconstructed with the B35f HeartView medium CaScore algorithm, generating a slice thickness of 3 mm, with 50% overlap between slices. For coronary CTA, iohexol (350 mg I/ml; Omnipaque, GE Healthcare, Stockholm, Sweden) was administered in an individual dose of 325 mg I per kg body weight and an injection time of 12 s. Five different acquisition protocols were used depending on body weight, heart rate and heart rate variability, described in detail elsewhere^20^.

### Clinical cohort

NCCT scans for calcium scoring were performed with 120 kV tube voltage, followed by coronary CTA after the administration, if needed, of intravenous metoprolol (0-30 mg) to reach a target heart rate of 60 bpm, and isosorbide dinitrate aerosol (1.25 mg) or sublingual nitrate (0.8 mg) to dilate the coronary arteries. Low-osmolal iodine contrast was used. Two hybrid PET-CT scanners with 64-row detectors were used (GE Discovery VCT and GE D690, General Electric Medical Systems, Waukesha, USA). Preferably, prospective image acquisition was used. Protocol details have been described previously^21^.

### EAT quantification

All non-contrast CT scans in the study were analyzed automatically for EATV and EATA with a fully automatic deep-learning-based model developed by our group^10^. Briefly, the model has been validated in a large series of cases from a sub-cohort technically identical to the SCAPIS cohort and found to generate segmentations in line with manual expert segmentations^15^. The model has a Dice-score for EATV at the upper practical limit of what has been both published on manual inter-reader agreement, and what our own inter-reader Dice-score has previously been^22^. The model automatically identifies potentially flawed segmentations/analyses, which can then be manually corrected if necessary. In the present study, all cases (828 population cases and 615 clinical cases) identified as potentially flawed were manually reviewed and corrected by an expert thoracic radiologist (DM) to represent anatomically accurate segmentations.

### Heart chamber segmentation

The heart segmentation network was developed using the state-of-the-art nnUNet framework^23^, trained on 200 CTA scans. Ground truth segmentation masks were first created by trained medical engineers and subsequently verified by a senior radiologist. To substantially reduce the manual effort required for volumetric annotation, the research software MiaLab^24^, which integrates interactive level-set segmentation with radial basis function (RBF) based 3D mesh generation and editing, was employed. Furthermore, an iterative annotation–training strategy was adopted: initial manual annotations from a limited dataset were used to train a preliminary model, which then assisted in producing additional labeled data for model refinement. The model outputs volumes of six cardiac structures: the left ventricular myocardium including the entire septum, the left ventricle, the right ventricle including its myocardium, the right atrium, the left atrium and the left atrial appendage. The sum of all output volumes is referred to as the “total heart volume” hereinafter.

### Statistical analysis

All statistical analysis was performed in R^25^, version 4.2.2. Statistical significance was defined as two-tailed p < 0.05. Test for normality was performed using visual examination of histograms and the Shapiro-Wilk test. Where continuous data was not normally distributed, the median with its interquartile range is reported. Categorical data is reported as percentages. The Mann-Whitney U-test was used to explore differences in continuous variables between groups, while the Fisher exact test was used to explore differences between groups for categorical variables.

Linear regression analysis was performed for the relationship between EATA and EATV, EATV and age and heart volume data. Normalization of EATV to patient height, body surface area (BSA) according to the formula of duBois & duBois, total ventricular (right + left ventricle), and total heart volumes was explored in the population cohort. Spearman’s rank correlation analysis was performed for EATA and EATV after these various normalization procedures. The relationship between EATVh and two metrics associated with increased cardiovascular risk, a Systematic Coronary Risk Evaluation 2 (SCORE2) of ≥ 7.5 10-year risk of atherosclerotic cardiovascular disease, and the presence of obstructive CAD defined as > 50% stenosis in any coronary artery, was assessed over quartiles of EATVh in the population cohort. In the clinical cohort SCORE2 was not available, and analysis was limited to significant stenosis. As a sensitivity analysis, logistic regression was performed in the population cohort to verify the association between raw and normalized EATV and the presence of obstructive CAD.

## Results

### Cohort characteristics

From the population cohort, a total of 12,409 men (49.3%) and 12,746 women (50.7%) with analyzable NCCT and CTA images were included. In the clinical cohort, a total of 1,039 men (41,9%) and 1,443 women (58.1%) with retrievable and analyzable NCCT and CTA images were included. The median age was comparable between sexes in the population cohort (57.3 years), whereas women in the clinical cohort were significantly older than men (65 versus 61 years) **[Table 1]**. BMI was slightly higher in the clinical (27.2 in men and 27.5 in women) than the population cohort (26.8 in men and 25.6 in women). The prevalence of diabetes, hypertension and dyslipidemia was overall markedly and comparably higher in the clinical cohort in both sexes. The proportion of current smokers, was higher among women in the population cohort than in the clinical cohort (12.3% vs. 8.9%) whereas it was slightly lower among men (12.3% vs. 17.1%).

**Table 1.** Basic clinical characteristics of the cohorts. Continuous numerical variables are shown as median values with their interquartile ranges in parentheses, while categorical variables are shown with their respective percentage and counts in parentheses. “A” denotes significance between sexes within each cohort, while “B” denotes significance between cohorts for same sex per the Mann-Whitney u-test for numericals and the Fisher exact test for categoricals.

| Variable | Population cohort (n=25,155) |  | Clinical cohort (n=2,482) |  |
| --- | --- | --- | --- | --- |
|  | Men (n=12,409) | Women (n=12,746) | Men (n=1,039) | Women (n=1,443) |
| Age [years] | 57.30<sup>B</sup> (53.50-61.10) | 57.30<sup>B</sup> (53.70-61.10) | 61.00<sup>AB</sup> (54.00-68.00) | 65.00<sup>AB</sup> (58.00-70.00) |
| BMI | 26.80<sup>AB</sup> (24.70-29.40) | 25.60<sup>AB</sup> (23.10-29.00) | 27.20<sup>B</sup> (24.90-30.15) | 27.50<sup>B</sup> (24.20-31.20) |
| EATV [ml] | 125.73<sup>AB</sup> (97.28-159.84) | 95.49<sup>AB</sup> (73.01-123.56) | 170.39<sup>AB</sup> (133.61-213.08) | 133.62<sup>AB</sup> (102.51-169.34) |
| EATA [HU] | -75.42<sup>AB</sup> (-78.25--72.63) | -74.79<sup>AB</sup> (-77.82--71.82) | -80.79<sup>AB</sup> (-83.55--77.65) | -79.79<sup>AB</sup> (-83.28--76.44) |
| BSA [m<sup>2</sup>] | 2.06<sup>AB</sup> (1.96-2.18) | 1.79<sup>A</sup> (1.69-1.89) | 2.05<sup>AB</sup> (1.93-2.19) | 1.79<sup>A</sup> (1.69-1.92) |
| Heart volume [ml] | 665.51<sup>AB</sup> (595.53-741.91) | 512.65<sup>AB</sup> (463.58-564.33) | 727.45<sup>AB</sup> (644.81-809.07) | 544.99<sup>AB</sup> (490.75-602.52) |
| EATV/BSA [ml/m<sup>2</sup>] | 60.85<sup>AB</sup> (48.32-75.51) | 53.42<sup>AB</sup> (41.93-67.22) | 81.88<sup>AB</sup> (66.81-101.50) | 74.57<sup>AB</sup> (58.96-92.86) |
| EATV/heart volume | 0.19<sup>AB</sup> (0.14-0.25) | 0.19<sup>AB</sup> (0.14-0.24) | 0.24<sup>AB</sup> (0.18-0.29) | 0.25<sup>AB</sup> (0.19-0.31) |
| Smoking (current) | 12.3%<sup>B</sup> (1525) | 12.3%<sup>B</sup> (1573) | 17.1%<sup>AB</sup> (178) | 8.9%<sup>AB</sup> (129) |
| Diabetes (any type) | 8.2%<sup>AB</sup> (1020) | 4.6%<sup>AB</sup> (586) | 18.1%<sup>AB</sup> (188) | 13.8%<sup>AB</sup> (199) |
| Hypertension | 19.4%<sup>AB</sup> (2408) | 16.9%<sup>AB</sup> (2159) | 58.1%<sup>B</sup> (604) | 57.5%<sup>B</sup> (830) |
| Dyslipidemia | 7.9%<sup>AB</sup> (985) | 5.4%<sup>AB</sup> (687) | 64.0%<sup>B</sup> (665) | 64.0%<sup>B</sup> (924) |
| Coronary stenosis > 50% | 8.6%<sup>AB</sup> (1063) | 2.3%<sup>AB</sup> (297) | 34.6%<sup>AB</sup> (360) | 15.9%<sup>AB</sup> (230) |

### Technical performance of the EAT analysis

From the 29,228 cases in the entire SCAPIS with available EAT data (of which the study cohort is a subset), 828 (2.83 %) segmentations were corrected manually after being flagged by the AI-based model as potentially flawed. A median absolute difference in EATV of 6.12 ml (IQR: 2.02-13.52) was observed between the automatic and manual segmentations, corresponding to an absolute difference of 6.68 % of the estimated EATV. Notably, the best three quartiles, i.e., 75% of corrected segmentations, had an absolute difference of less than 10.95 %, and the best quartile less than 2.06 %. Only the worst quartile comprising 207, i.e., 0.71 % of all segmented cases had an error of more than 10.95%.

Among clinical patients, 615 (19.19%) cases from a total of 3,205 with available EAT data (of which the study cohort of 2,482 is a subset) were flagged and corrected. A median absolute difference in EATV of 18.67 ml (IQR: 8.28-37.36) was observed, corresponding to an absolute difference of 11.44 % of the estimated EATV. The best three quartiles, i.e. 75% of corrected segmentations, had an absolute difference of less than 20.97%. The best quartile had an absolute difference of less than 4.57 %, comparable to results in the population cohort.

### Technical performance of the heart volume analysis

In total, 1,445 (5.7 %) cases in the population cohort and 3,037 (100%) clinical cases with available heart chamber segmentations, of which the study cohort is a subset, were manually reviewed by an expert thoracic radiologist (DM). From these, in the clinical cohort, 43 cases (1.42 %) had varying degrees of imperfections not deemed to influence total heart volume estimates and 7 were deemed to be failed with respect to anatomical segmentation (0.23 % of all analyses), most of them due to the absence of contrast in the images due to timing or technical errors. These failed segmentations were not removed in the primary analyses, since they would only minimally influence results, and we wanted to test performance in a real-world cohort under realistic conditions. Among the manually reviewed cases in the population cohort, 5 segmentations had varying degrees of imperfections not deemed to influence total heart volume estimates (0.35 % of all analyses), and none were deemed to be failed (0.00 %). In some of the population cases, the heart was incompletely imaged, with substantial parts missing, typically inferiorly. Such cases were excluded in the analyses, based on a requirement of minimum 40 ml of left ventricular myocardium having to be segmented (10 cases in total).

### EAT volumes, attenuation and normalization procedures

Absolute EATV in the population cohort was not normally distributed, and showed significant differences between sexes, with men having higher amounts of EAT across the entire age range. The median value for the entire population cohort (109.7 ml) was comparable to what has been previously reported in a selection of studies based on manual or semi-automatic quantification **[Supplement: Table S.1]**, all of which have a weighted mean of 104.4 ml. EATV increased slightly with age in both sexes **[Figure 1]** and **[Supplement: Table S.2]**. In both sexes, there was a clear inverse correlation between EATA and EATV, which was tightened upon normalization, with less scattering around the regression line, most notably with normalization to heart volume **[Figure 2]**. There was only a minimal decrease in heart volumes with age in both men and women **[Supplement: Figure S.3]**.

**Figure 1.**
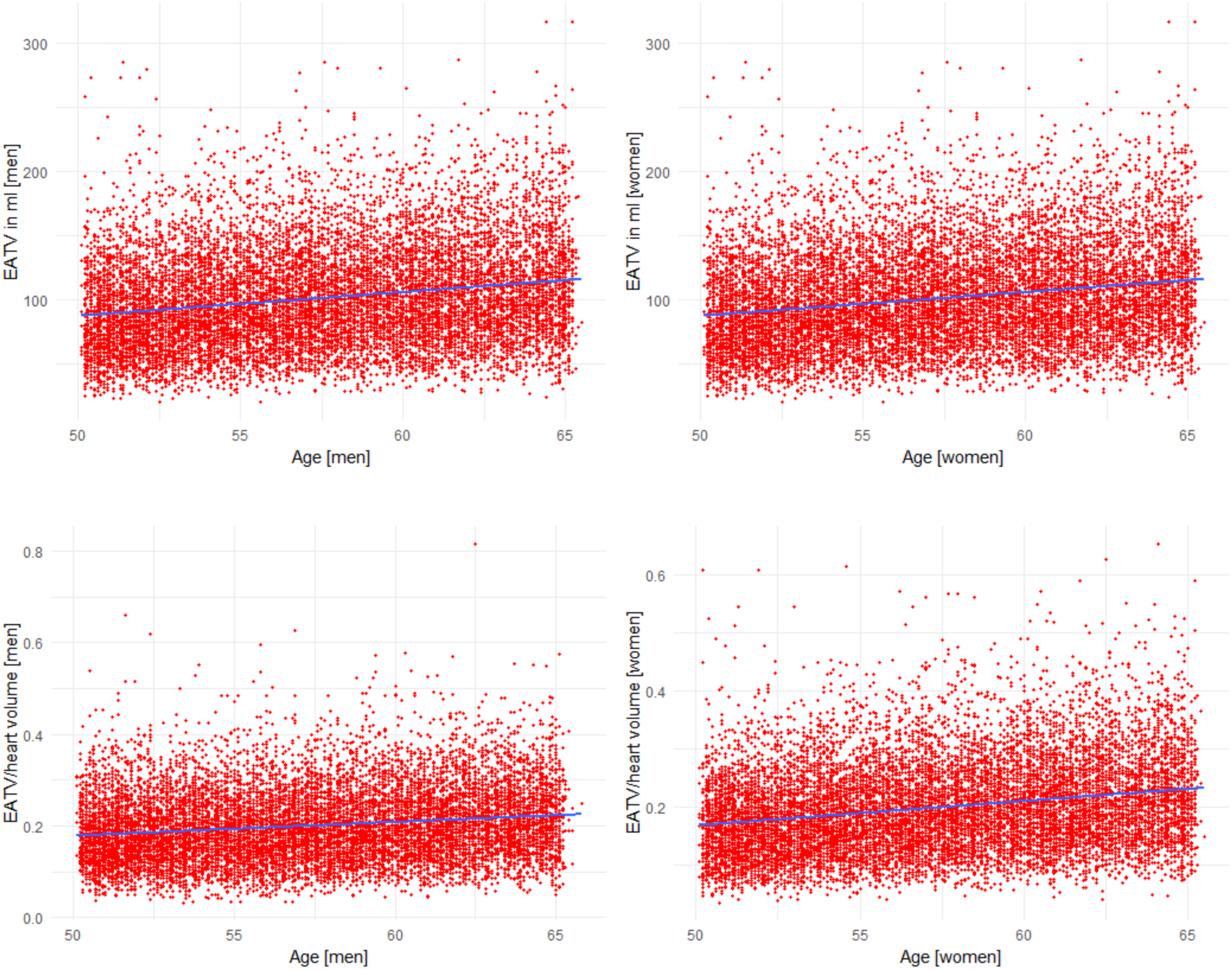
Relationship between EATV and age in men (n=12,409) and women (n=12,746) in the population cohort. The upper row shows non-normalized EATV and the lower row shows EATV normalized to total heart volume.

**Figure 2.**
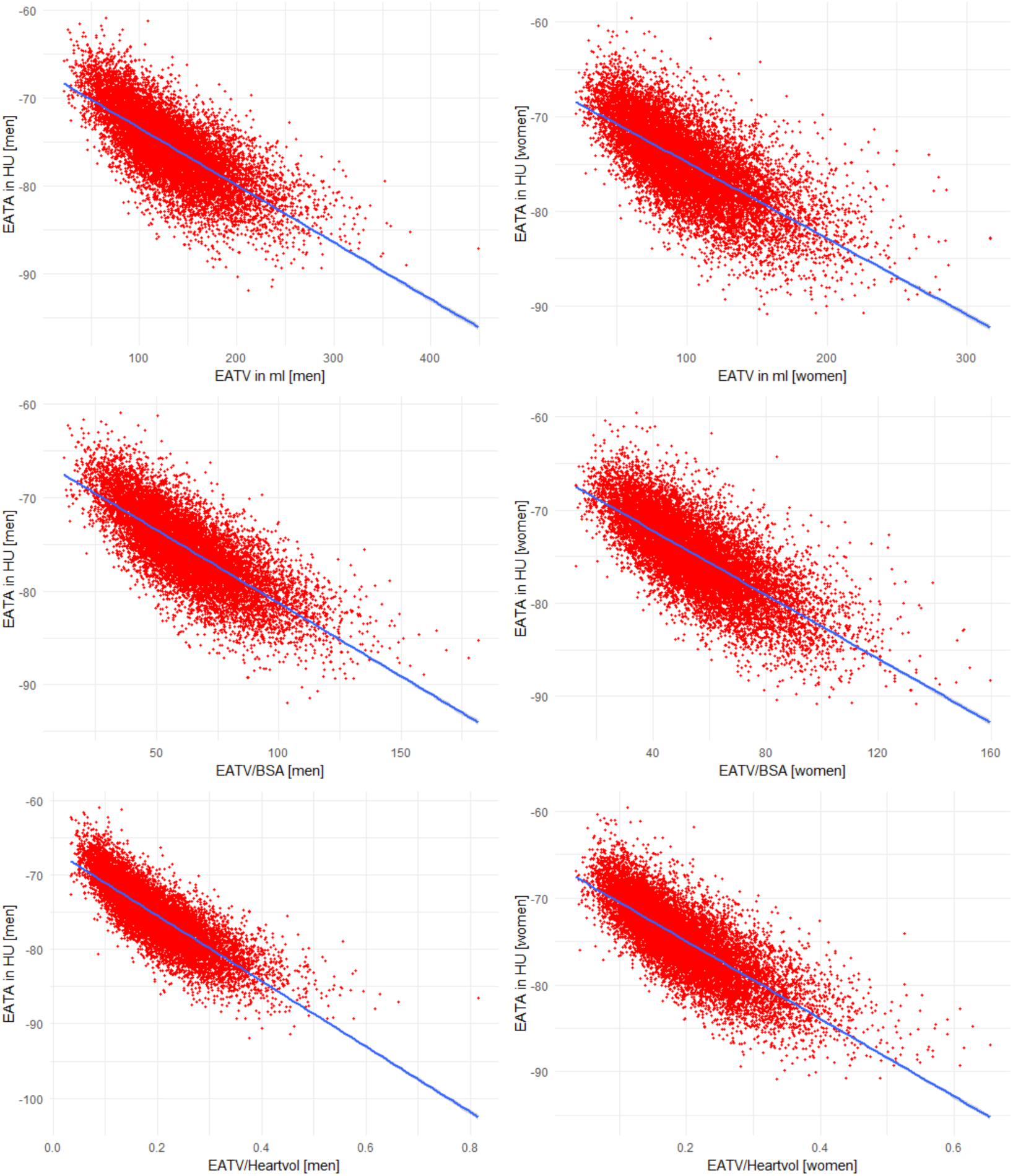
Epicardial adipose tissue attenuation (EATA) as a function of epicardial adipose tissue volume (EATV) in men (n=12,409) and women (n=12,746) in the population cohort. The upper row shows non-normalized EATV, the mid row EATV normalized to body surface area, and the lower row EATV normalized to heart volume.

In the population cohort, for both sexes alike, linear regression coefficients per quartile increase in the underlying EATV metric were very similar, translating into a decrease in EATA for every quartile of increase of approximately 4.1 HU for non-normalized EATV, and 4.5 HU for EATV normalized to heart volumes **[Table 2a]**.

**Table 2a.**
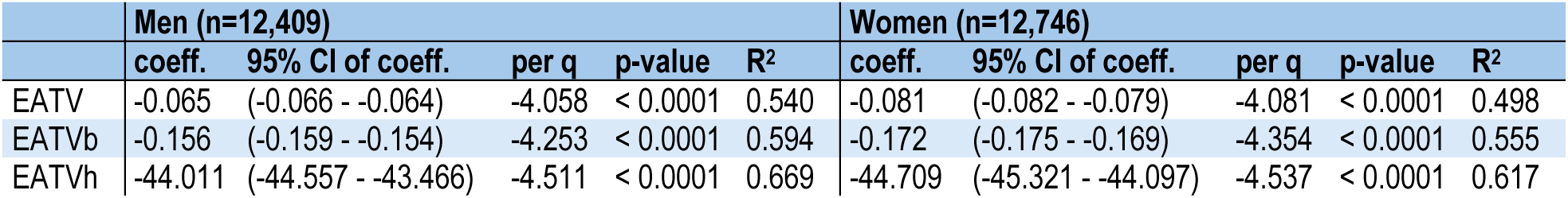
Linear regression of epicardial adipose tissue attenuation as a function of epicardial adipose tissue volumes in the population cohort after normalization procedures. The regression coefficient, its 95% confidence interval, the regression coefficient per quartile of increase in EATV, the p-value and the R^2^-value are shown in the columns by sex. The top row shows non-normalized EATV, the mid row EATV normalized to BSA and the bottom row EATV normalized to heart volume.

Normalization of EATV to height, BSA, total ventricular, and total heart volumes all saw improved rank correlation between EATA and EATV **[Supplement: Table S.2b]**, and in both cohorts, normalization to total heart volume was superior in both sexes and yielded similar improvement **[Table 2b]**. Notably, the linear regression modeling in the population cohort showed significant improvement in R^2^ with normalization to heart volume in both sexes, which was considered testimony to better model fit.

**Table 2b.** Spearman’s rank correlation analyses between epicardial adipose tissue volumes and epicardial adipose tissue attenuation after various normalization procedures (“EATVn”) in the population and clinical cohorts. The uppermost row shows non-normalized data, the second row EATV normalized to body surface area, the last row EATV normalized to total heart volume (all chambers).

|  | Population cohort |  |  |  |  | Clinical cohort |  |  |  |
| --- | --- | --- | --- | --- | --- | --- | --- | --- | --- |
|  | Men |  | Women |  |  | Men |  | Women |  |
| EATVn × EATA | Corr. | p-value | Corr. | p-value | EATVn × EATA | Corr. | p-value | Corr. | p-value |
| EATV | -0.747 | NA | -0.721 | NA | EATV | -0.707 | NA | -0.723 | NA |
| EATV/BSA | -0.776 | < 0.0001 | -0.752 | < 0.0001 | EATV/BSA | -0.730 | < 0.0001 | -0.755 | < 0.0001 |
| EATV/total heart vol. | -0.832 | < 0.0001 | -0.796 | < 0.0001 | EATV/total heart vol. | -0.800 | < 0.0001 | -0.796 | < 0.0001 |

The basic relationships between EATA and EATV were seen also in the clinical cohort, despite the absolute EATV being substantively higher **[Tables 5 and 6, Supplement: Figures S.1 and S.2]**.

### General population normal values for EAT

In the population cohort, EATV normalized to total heart volume (EATVn), was nearly identical between sexes, with a subtle, but continuous increase with age **[Supplement: Table S.2] [Figure 1]**. Pooled results showed a more marked stepwise increase with age across 5-year age strata. This trend could be replicated in the clinical cohort **[Table 3]**, where EATV normalized to heart volume showed very good agreement between sexes, while raw non-normalized EATV differed considerably. EATA showed less pronounced differences by sex, especially among the older individuals in the population cohort (60-65 years), where median values were similar. This was seen also in the clinical cohort **[Table 4]**.

**Table 3.** Epicardial adipose tissue volumes by 5-year age strata in men and women in the population and the clinical cohorts. The top table shows non-normalized EATV, the mid table shows EATV normalized to body surface area and the bottom table shows EATV normalized to heart volume.

| Population cohort |  |  |  |  |  |  | Clinical cohort |  |  |  |  |  |  |
| --- | --- | --- | --- | --- | --- | --- | --- | --- | --- | --- | --- | --- | --- |
|  | Men |  |  | Women |  |  |  | Men |  |  | Women |  |  |
| Age | n | EATV | IQR | n | EATV | IQR | Age | n | EATV | IQR | n | EATV | IQR |
| 50-54 | 3532 | 116.8 | 90.7-150.1 | 3556 | 84.9 | 64.6-110.7 | 50-54 | 264 | 153.3 | 119.2-190.2 | 224 | 108.8 | 83.4-141.8 |
| 55-59 | 4111 | 124.0 | 96.1-157.3 | 4277 | 95.8 | 73.7-122.7 | 55-59 | 172 | 167.4 | 132.3-211.3 | 203 | 131.8 | 95.4-164.5 |
| 60+ | 4766 | 134.1 | 104.3-169.4 | 4913 | 103.5 | 80.1-133.0 | 60+ | 603 | 174.0 | 139.2-223.1 | 1016 | 140.5 | 107.9-177.9 |

EATV – normalized to BSA [ml/m2]
| Population cohort |  |  |  |  |  |  | Clinical cohort |  |  |  |  |  |  |
| --- | --- | --- | --- | --- | --- | --- | --- | --- | --- | --- | --- | --- | --- |
|  | Men |  |  | Women |  |  |  | Men |  |  | Women |  |  |
| Age | n | EATV | IQR | n | EATV | IQR | Age | n | EATV | IQR | n | EATV | IQR |
| 50-54 | 3532 | 56.195 | 44.73-70.36 | 3556 | 47.441 | 36.99-59.72 | 50-54 | 264 | 73.283 | 57.97-87.92 | 224 | 59.532 | 46.01-74.95 |
| 55-59 | 4111 | 59.675 | 47.55-73.88 | 4277 | 53.449 | 42.09-66.53 | 55-59 | 172 | 81.444 | 66.23-101.09 | 203 | 72.392 | 56.58-89.11 |
| 60+ | 4766 | 65.506 | 51.84-80.20 | 4913 | 58.243 | 46.17-72.55 | 60+ | 603 | 86.142 | 71.10-105.83 | 1016 | 77.805 | 63.13-95.25 |

EATV – normalized to heart volume [ml/ml]
| Population cohort |  |  |  |  |  |  | Clinical cohort |  |  |  |  |  |  |
| --- | --- | --- | --- | --- | --- | --- | --- | --- | --- | --- | --- | --- | --- |
|  | Men |  |  | Women |  |  |  | Men |  |  | Women |  |  |
| Age | n | EATV | IQR | n | EATV | IQR | Age | n | EATV | IQR | n | EATV | IQR |
| 50-54 | 3532 | 0.175 | 0.13-0.23 | 3556 | 0.166 | 0.12-0.21 | 50-54 | 264 | 0.206 | 0.16-0.27 | 224 | 0.205 | 0.15-0.26 |
| 55-59 | 4111 | 0.187 | 0.14-0.24 | 4277 | 0.187 | 0.14-0.24 | 55-59 | 172 | 0.238 | 0.18-0.29 | 203 | 0.243 | 0.18-0.30 |
| 60+ | 4766 | 0.205 | 0.16-0.26 | 4913 | 0.207 | 0.16-0.26 | 60+ | 603 | 0.245 | 0.19-0.31 | 1016 | 0.256 | 0.20-0.32 |

**Table 4.** Epicardial adipose tissue attenuation by 5-year age strata in men and women in the population and clinical cohorts.

| Population cohort |  |  |  |  |  |  | Clinical cohort |  |  |  |  |  |  |
| --- | --- | --- | --- | --- | --- | --- | --- | --- | --- | --- | --- | --- | --- |
|  | Men |  |  | Women |  |  |  | Men |  |  | Women |  |  |
| Age | n | EATA | IQR | n | EATA | IQR | Age | n | EATA | IQR | n | EATA | IQR |
| 50-54 | 3532 | -75.0 | -77.7--72.3 | 3556 | -73.7 | -76.6--70.8 | 50-54 | 264 | -80.0 | -82.9--77.3 | 224 | -77.3 | -80.5--73.2 |
| 55-59 | 4111 | -75.4 | -78.1--72.6 | 4277 | -74.7 | -77.7--71.8 | 55-59 | 172 | -81.0 | -84.4--78.0 | 203 | -79.2 | -83.5--75.8 |
| 60+ | 4766 | -75.8 | -78.8--73.0 | 4913 | -75.7 | -78.6--72.6 | 60+ | 603 | -80.9 | -83.7--77.8 | 1016 | -80.4 | -83.7--77.2 |

### EATV in relation to obstructive CAD and high-risk by SCORE2 in the general population

Both the prevalence of obstructive CAD and SCORE2 of ≥ 7.5 increased consistently over quartiles of EATVh in both sexes (p < 0.001 for both) **[Figure 3]**.

**Figure 3.**
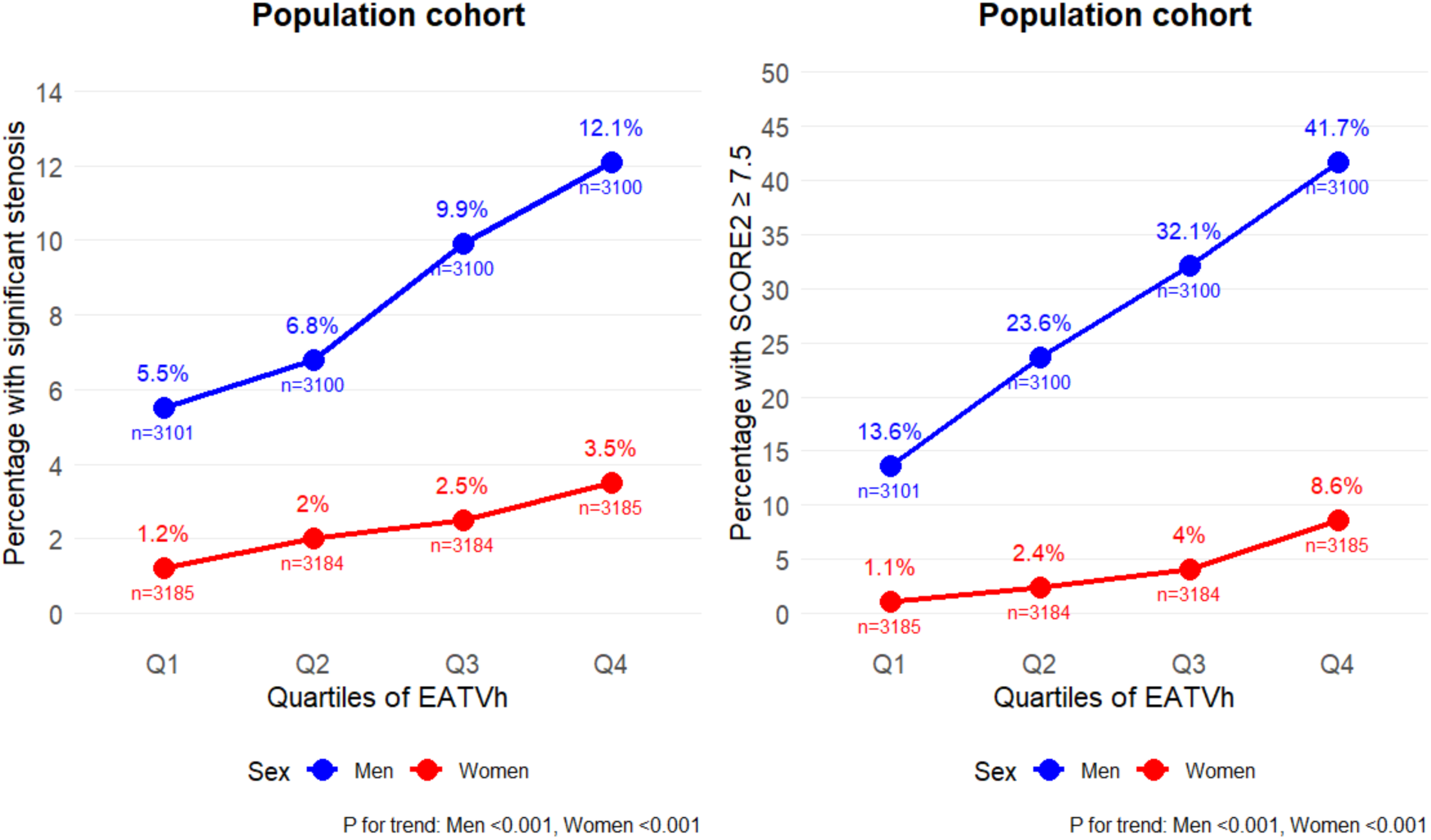
The prevalence of obstructive coronary artery disease (defined as stenosis of > 50 % in any coronary artery) and high cardiovascular risk (defined as SCORE2 ≥ 7.5) by quartiles of epicardial adipose tissue normalized to heart volume (EATVh) in the population cohort (n=25,155). P-values for the trend over quartiles is shown beneath each plot.

Logistic regression in the population cohort with obstructive CAD as a binary outcome measure, and both EATV and EATV normalized to BSA and heart volume as dependent variables yielded similar OR in men and women per quartile increase in the underlying variables (p-value < 0.001 for all) **[Supplement: Table S.3]**. This was was seen regardless of normalization procedure. The increase in the odds of having obstructive CAD was somewhat more marked in women (61.9%) than in men (47.7%) for every quartile increase in EATV normalized to heart volume. After adjustment for clinical risk factors, this difference was attenuated. Among individuals having obstructive CAD, the median EATVh was significantly higher irrespective of which age stratum they belonged to **[Figure 4]**.

**Figure 4.**
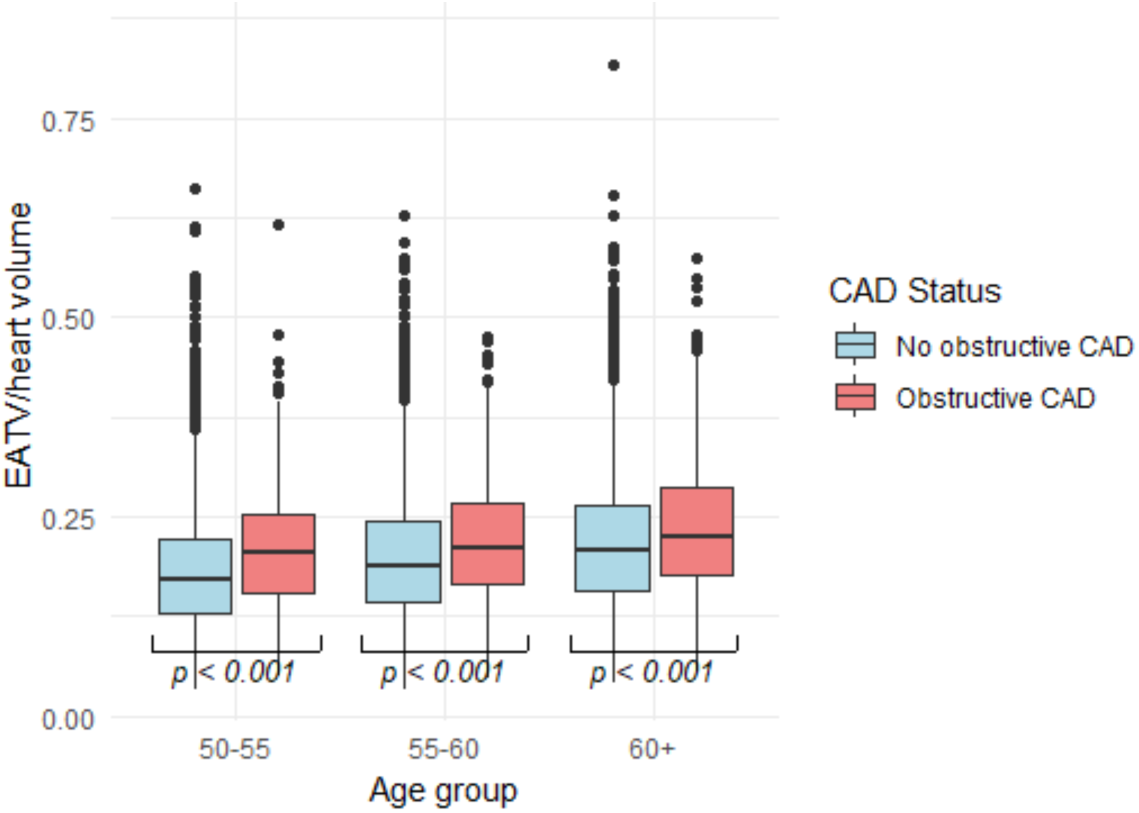
Box-plot showing the distribution of EATV normalized to heart volume over age strata among participants in the population cohort (n=25,155) grouped by the presence or absence of obstructive coronary artery disease defined as stenosis of > 50 % in any coronary artery. The p-values for the Mann-Whitney u-test between the groups are shown below each pair.

### EATV in relation to obstructive CAD in the clinical cohort

The prevalence of obstructive CAD increased over quartiles of EATVh in both sexes (p < 0.05) **[Supplement: Figure S.4]** despite higher overall levels, reaching 41% of men and 19% of women in the fourth EATVh quartile (up from 25% and 13% respectively in the first quartile). This is to be compared to 12% in men and 3.5% in women in the fourth quartile in the population cohort.

### Suggested clinical reference values for EATVh

A possible general cut-off such, that 95% of the cases with obstructive CAD would have an EATVh above it, was calculated for the entire population cohort **[Table 5]**. Notably, this cut-off is somewhat lower, but close to the lower first quartile values by age **[Table 3]**. Since EATVh is practically sex-independent and trends were similar between sexes and also cohorts, we decided to use one single cut-off. For reasons of simplicity, at this stage, an age-independent cut-off of 0.10 in EATVh was chosen, since variations were comparatively small across age strata of the prevalence of CAD. In addition, quartile based cut-offs were also explored, corresponding to an EATVh of 0.14 (approximate first quartile upper limit) and 0.25 (approximate fourth quartile lower limit) **[Supplement: Table S.4]**. In the population cohort, the low cut-off of 0.10 had a negative predictive value for obstructive CAD of 95% in men and 99.2% in women (overall, both sexes: 97.1%), with a negative likelihood ratio ((-)LR) of 0.56 and 0.34 respectively, indicating better performance in women. For high-risk by SCORE2, the negative predictive value was 90.6% and 99.3% respectively, with (-)LR:s of 0.27 and 0.17. Expectedly, metrics deteriorated somewhat when increasing the cut-off to 0.14, although in women the NPV for high-risk by SCORE2 was still high, with a low (-)LR. Finally, the high cut-off of 0.25 came with low positive predictive values, but moderate (+)LR for high-risk by SCORE2 in particular.

**Table 5.** Proposed cut-off in EATV normalized to heart volumes (EATVh) based on 95% of cases in the population cohort with obstructive CAD having higher EATVh.

| Age group | n | cut-off | cases above [%] |
| --- | --- | --- | --- |
| 50-54 | 254 | 0.0974 | 94.9 |
| 55-59 | 424 | 0.104 | 94.8 |
| 60+ | 682 | 0.116 | 94.9 |

In the clinical cohort, the low cut-off of EATVh 0.10 yielded an NPV for obstructive CAD of 82.6% in men and 93.5% in women (overall, both sexes: 88.9%), with (-)LR:s of 0.40 and 0.36 respectively **[Supplement: Table S.5]**. Increasing the cut-off to 0.14 in EATVh still would give an NPV of 83.2% and 89.9% for men and women, with (-)LR increasing to 0.38 and 0.59, indicating less utility as a rule-out on its own merits.

## Discussion

In this multi-center study comprising more than 27,500 individuals with comprehensive NCCT and CTA data we explored EAT volumes and attenuation with the aims of defining normal reference values in the general population and to compare them with values from a selected symptomatic population with suspected CAD. Our findings suggest that EATV should be normalized, whenever possible, to mitigate the effects of body size, and that normalization to total heart volume is preferable. Sex differences, which are pronounced in non-normalized EATV, while somewhat less so in EATV normalized to BSA, are almost entirely neutralized in EATV normalized to total heart volume. Furthermore, the correlation between EATA and EATV is significantly improved when normalization to total heart volume is performed, and this normalization of EATV did not negatively influence the prediction of obstructive CAD in uni- or multivariable logistic regression. A subtle but important increase in EATV with age in both sexes should also be acknowledged, since it can be an important confounder in EAT analyses.

Until now, reliable reference values for EATV and EATA have been largely unavailable, limiting the use of EAT in both research and as a potential clinical tool for risk stratification. It has been known from at least one study^13^, that EATV increases with age, but this effect has not been thoroughly described in any larger cohort reflecting the true background population. It has been clear from previous studies that EATA is inversely correlated to EATV^26–28^. We have earlier shown, in a study on nearly 2,000 individuals, focusing on glucose disorders covering the range from normoglycemia to T2-diabetes, that the relationship is very tight, and that EATA might be a predictor of impaired glucose tolerance, and thus, early glucose disorders^15^. The logic behind the strong interrelation can be explained by the expansion in EATV primarily being driven by the expansion of adipocytes through the storage of lipids^17,19^. The resulting increased lipid content of the EAT will inevitably lead to a lower mean radiodensity and EATA, given that all other influencing factors are equal. Potentially, inflammatory changes in the EAT might affect the EATA in the contrary direction but will be unlikely to affect the total EAT volume, presumed that they are located in a small volume around the coronary arteries^29–34^. Consequently, Eisenberg et al. found that a lower EATA was a predictor of coronary events^35^.

The question of normalization of EATV to anthropometric measures has been scarcely addressed until now, with several studies having used normalized EATV to BSA, but without exploring the relationships closer, and with no convincing experimental data supporting the choice. It is clear from our results, that EATV is strongly influenced by body size, also explaining to large parts the significant sex differences in EATV both in our cohort mirroring a normal background population and our clinical cohort.

It is well established, almost axiomatic, that men at an average are not only taller and heavier, but also have comparably more voluminous hearts^36^. Not taking this into account would presumably reduce the precision of EATV as a factor for finding obese individuals, and individuals with truly abnormally high amounts of EATV, which plausibly are the ones at risk of experiencing coronary events. It is self-explanatory from the geometric relationships, that a very tall, but lean man, with a thin rim of EAT around a big heart, could have the same absolute EATV measured as a short, but obese man with abnormally increased EATV, sporting a thicker layer of EAT around a smaller heart. In our analyses, the total heart volume, measured as the volume of all chambers, gave the best correlation between EATA and EATV, and thus, theoretically, the most correct approximation of the true, body-size adjusted EATV. It should be noted that total heart volume varies relatively little over the heart cycle^37^ and therefore should be a more robust metric than ventricle volumes in a setting, such as ours, where CTA images are not always acquired in the same phase of the heart cycle.

A possible drawback of using the total heart volume might be encountered in patients with manifest heart disease, which are prone to cardiomegaly, and therefore might have normalized EATV which does not correctly reflect their level of EAT expansion. This, however, did not limit the usefulness of heart volume in normalization in the clinical cohort, where the prevalence of CAD was relatively high. Furthermore, we believe that the greatest advantage of normalization comes into fruition when screening individuals who have no previously diagnosed heart disease and would thus not be affected by cardiomegaly to any significant extent. To be able to reliably utilize EATV as a tool for risk assessment, i.e., in conjunction with calcium scoring, normalization to heart volumes would be preferable. Our model for heart chamber segmentation is currently being adapted to handle NCCT images, and EATV assessment together with normalization should therefore be available in a “one-stop-shop” setting for calcium-scoring images, once the method of measuring EATV is incorporated into clinical routine use.

In line with previous studies^3,38–40^, across age strata in the population cohort EATV normalized to heart volume (EATVh) was higher among individuals having obstructive CAD. There was also a significant trend in both the population and the clinical cohorts of increasing prevalence of obstructive CAD over quartiles of EATVh. To explore the potential clinical usefulness of a cut-off in EATVh, a level above which 95% of obstructive CAD cases are included, was calculated, being only slightly lower than the lower first quartile of EATVh by age. To simplify applicability, we decided to approximate this to EATVh of 0.1, which, when applied, yielded a high negative predictive value (up to 93.5% in women) also in the clinical cohort, still with fairly low negative likelihood ratio.

It can always be argued, that the cut-offs should be differentiated by sex, age and also, possibly adjusted to the pre-test probability. However, this was outside of the scope and means of the present study, where we didn’t have access to SCORE2 in the clinical cohort, where we also had a limited set of clinical data available. Still, we could demonstrate that cut-offs in EATVh derived from a population cohort with low CAD burden are transferable to a clinical cohort with considerably higher disease burden. In our opinion, this opens for a broader utility of EATVh in risk stratification, especially as an auxiliary metric to be used in addition to established and existing metrics such as SCORE2. Exploration of EATVh as a risk-marker, both alone and in combination with other image-derived markers such as the coronary artery calcium score, especially with respect to long-term outcome, is the topic for further research, preferably with validation over a broad spectrum of disease manifestations.

### Strengths and limitations of the study

The major strength lies in the large cohorts with high-quality image data available, which represent the most extensive series of individuals examined to date with automated EAT and heart chamber quantification. This has allowed for establishing normal reference values by age and sex in a Northern European background population and validation in a clinical cohort evaluated for chest pain. The homogenous nature of the study population would purportedly limit generalizability to broader populations with other or mixed ethnicity, and although the exact variability over ethnic groups in EAT and heart volumes would not be expected to be very high, at least with respect to normalized values, this naturally must be addressed in future studies.

Potentially, automated, AI-based image analysis always carries the risk of errors, which can be hard to find or even understand; this is an inherent limitation of all work performed without direct human supervision. However, our previously carried out validation work, further corroborated by the extensive manual reviewing of potentially flawed segmentations carried out in this study, has demonstrated that automated analyses can achieve the same degree of accuracy as manual measurements, at least at a group level comprising hundreds of individual cases. Also, notably, our model has been shown to be robust enough to translate well to a clinical cohort, materially different in technical image parameters than the cohort it was initially developed for. This, we believe, increases the chances that further cohorts can be successfully analyzed with the model, perhaps resolving the question on generalizability over ethnic groups, and also a wider age span, than what was available in our study. While the number of flawed segmentations in the clinical cohort were higher, due to images of more inconsistent quality and a significant proportion of patients being equipped with pacemakers, resulting in a significantly increased number of manually corrected images, the best quartile among the corrected were of comparable segmentation quality to the best quartile in the population cohort. Finally, it deserves mentioning, that automated analysis is a prerequisite for the analysis of any large cohort study, and likely also a critical facilitator for the clinical implementation of EAT analysis.

## Conclusions

In this study, which allowed for granular tabulation of normal values of EAT volumes and attenuation by age and sex in a true Northern European population sample, we could demonstrate significant sex differences in EAT volumes and that these can be canceled by normalizing EATV to total heart volumes. Validation in a clinical cohort evaluated for chest pain and with different technical image data confirmed the utility of the concept, and a potential clinically useful cut-off for normalized EATV was proposed based on the occurrence of obstructive coronary artery disease. This cut-off had an overall negative predictive value for obstructive CAD of 97.1% in the general population and of 88.9% in the population evaluated for chest pain.

## Acknowledgments

This research was conducted using the Swedish Cardiopulmonary Bioimage Study (SCAPIS) Resource under Petition 1086. The Swedish CArdioPulmonary bioImage Study (SCAPIS) is funded by the Swedish Heart-Lung Foundation. Additional support is provided by the Knut and Alice Wallenberg Foundation, the Swedish Research Council, VINNOVA (Sweden’s Innovation agency), the participating Universities and University Hospitals. The Finnish part of the study was funded by the Finnish Foundation for Cardiovascular Research, Finnish State Research Funding, and the Research Council of Finland. Dr. Molnar’s work has been co-funded by the European Union’s Horizon Europe Framework program for research and innovation 2021-2027 under the Marie Slodowska-Curie grant agreement No. 101126611.

Additional funding for Dr. Molnar’s work has been provided by the Swedish Medical Society, the Swedish Heart-Lung Foundation, the Swedish Heart Union and the Gothenburg Medical Society.

## Data availability

Data used in this study can be made available upon reasonable request to the authors of the paper.

---

**Supplement: Table S.1.**
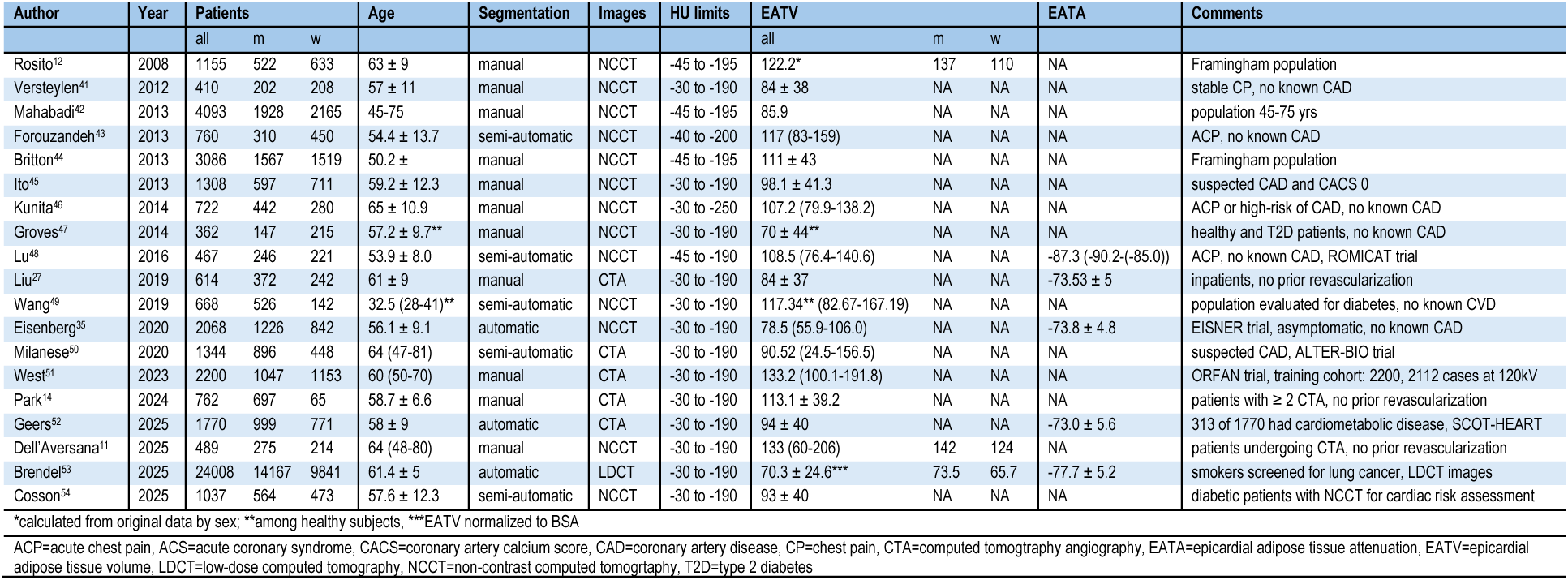
Selection of methodologically or demographically important computed tomography-based studies quantifying EAT in the literature. Numbers are median with interquartile range in parentheses or mean with ± SD.

**Supplement: Table S.2.**
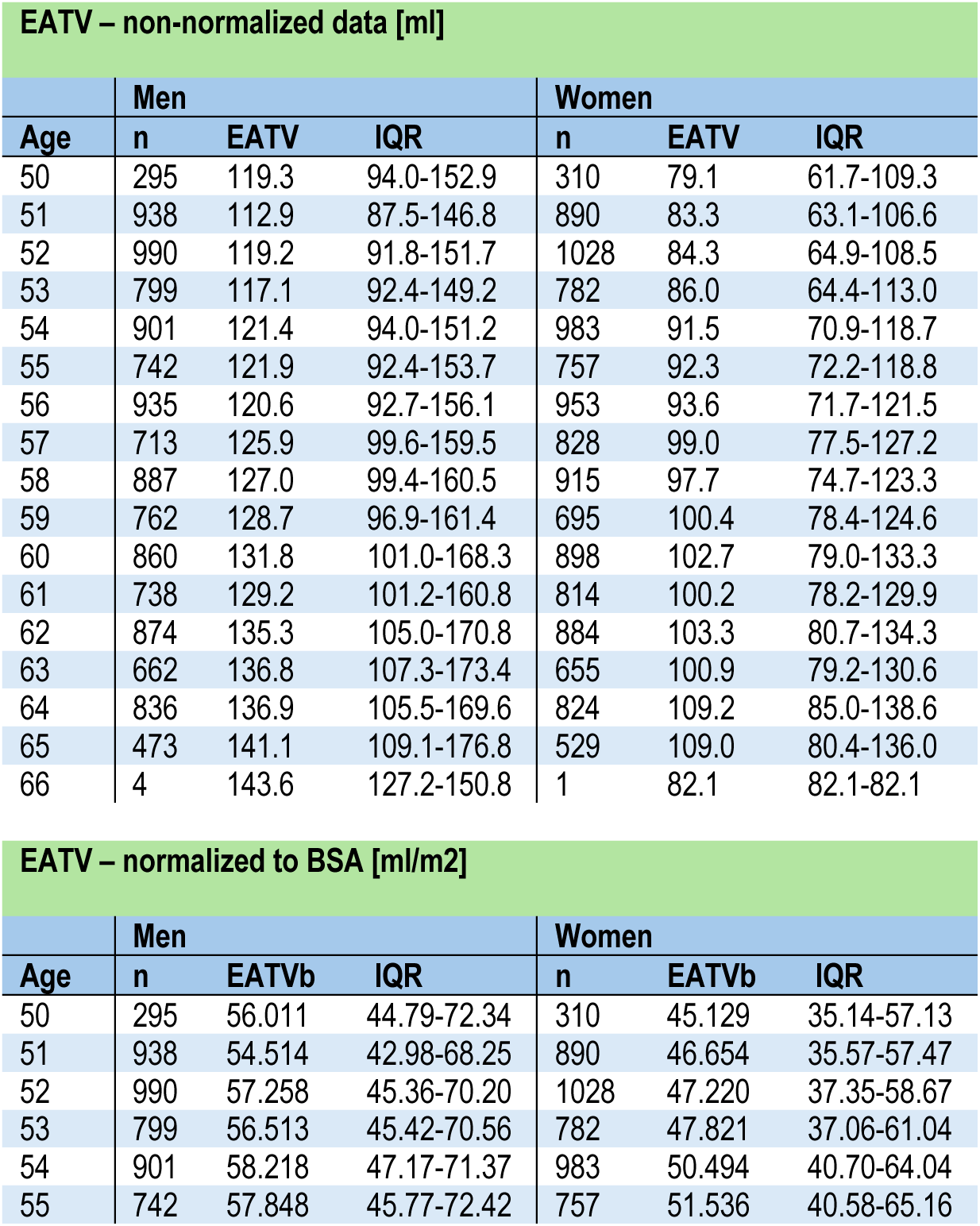
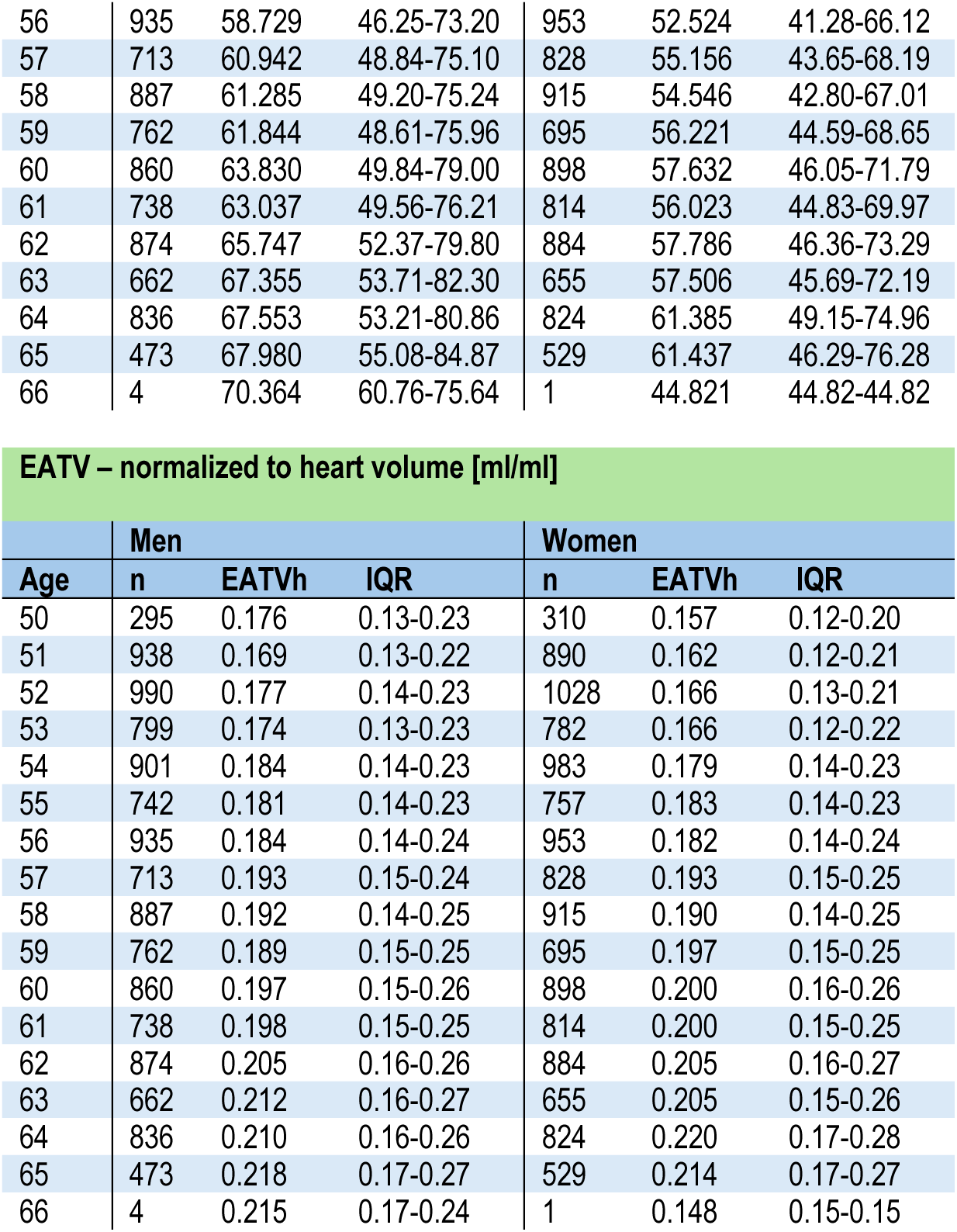
Epicardial adipose tissue volumes by age (50 to 65 years) in men and women in the population cohort. The top table shows non-normalized EATV, the mid table shows EATV normalized to body surface area and the bottom table shows EATV normalized to heart volume.

**Supplement: Table S.2b.** Spearman’s rank correlation analyses between epicardial adipose tissue volumes and epicardial adipose tissue attenuation after various normalization procedures (“EATVn”) in the population cohort. The uppermost row shows non-normalized data, the second row EATV normalized to body surface area, the last row EATV normalized to total heart volume (all chambers).

| Population cohort |  |  |  |  |
| --- | --- | --- | --- | --- |
|  | Men |  | Women |  |
| EATVn × EATA | Corr. | p-value | Corr. | p-value |
| EATV | -0.747 | NA | -0.721 | NA |
| EATV/height | -0.761 | < 0.0001 | -0.735 | < 0.0001 |
| EATV/BSA | -0.776 | < 0.0001 | -0.752 | < 0.0001 |
| EATV/LV+RV vol. | -0.809 | < 0.0001 | -0.772 | < 0.0001 |
| EATV/total heart vol. | -0.832 | < 0.0001 | -0.796 | < 0.0001 |

**Supplement: Table S.3.** Logistic regression analyses in the population cohort with the presence of obstructive coronary artery disease defined as > 50 % stenosis in any coronary vessel as outcome and non-normalized EATV, EATV normalized to body surface area (EATVb) and EATV normalized to heart volume (EATVh) as predictors in univariable modelling, and with the addition of clinical risk factors in multivariable modelling. Odds ratios and their respective confidence intervals are shown for the outcome per quartile increase in the underlying predictor.

| Outcome: presence of obstructive coronary artery disease |  |  |  |  |  |  |
| --- | --- | --- | --- | --- | --- | --- |
| Predictor | Model | Sex | OR | CI 95 | p-value | n |
| EATV | Unadjusted | Men | 1.467 | (1.358 - 1.585) | < 0.0001 | 12401 |
| EATV | + Smoking | Men | 1.409 | (1.303 - 1.524) | < 0.0001 | 12401 |
| EATV | + Smoking + Diabetes | Men | 1.330 | (1.227 - 1.442) | < 0.0001 | 12386 |
| EATV | + Smoking + Diabetes + Hypertension | Men | 1.245 | (1.146 - 1.352) | < 0.0001 | 12386 |
| EATVb | Unadjusted | Men | 1.539 | (1.424 - 1.663) | < 0.0001 | 12401 |
| EATVb | + Smoking | Men | 1.474 | (1.362 - 1.596) | < 0.0001 | 12401 |
| EATVb | + Smoking + Diabetes | Men | 1.394 | (1.285 - 1.512) | < 0.0001 | 12386 |
| EATVb | + Smoking + Diabetes + Hypertension | Men | 1.312 | (1.207 - 1.426) | < 0.0001 | 12386 |
| EATVh | Unadjusted | Men | 1.477 | (1.369 - 1.594) | < 0.0001 | 12401 |
| EATVh | + Smoking | Men | 1.411 | (1.304 - 1.525) | < 0.0001 | 12401 |
| EATVh | + Smoking + Diabetes | Men | 1.325 | (1.223 - 1.437) | < 0.0001 | 12386 |
| EATVh | + Smoking + Diabetes + Hypertension | Men | 1.259 | (1.16 - 1.367) | < 0.0001 | 12386 |
| EATV | Unadjusted | Women | 1.620 | (1.421 - 1.846) | < 0.0001 | 12738 |
| EATV | + Smoking | Women | 1.536 | (1.343 - 1.756) | < 0.0001 | 12738 |
| EATV | + Smoking + Diabetes | Women | 1.428 | (1.243 - 1.641) | < 0.0001 | 12730 |
| EATV | + Smoking + Diabetes + Hypertension | Women | 1.303 | (1.13 - 1.503) | < 0.0001 | 12730 |
| EATVb | Unadjusted | Women | 1.698 | (1.484 - 1.942) | < 0.0001 | 12738 |
| EATVb | + Smoking | Women | 1.599 | (1.394 - 1.836) | < 0.0001 | 12738 |
| EATVb | + Smoking + Diabetes | Women | 1.496 | (1.299 - 1.724) | < 0.0001 | 12730 |
| EATVb | + Smoking + Diabetes + Hypertension | Women | 1.378 | (1.192 - 1.594) | < 0.0001 | 12730 |
| EATVh | Unadjusted | Women | 1.619 | (1.424 - 1.841) | < 0.0001 | 12738 |
| EATVh | + Smoking | Women | 1.529 | (1.34 - 1.745) | < 0.0001 | 12738 |
| EATVh | + Smoking + Diabetes | Women | 1.419 | (1.237 - 1.627) | < 0.0001 | 12730 |
| EATVh | + Smoking + Diabetes + Hypertension | Women | 1.324 | (1.151 - 1.524) | < 0.0001 | 12730 |

**Supplement: Table S.4.** Diagnostic performance in the population cohort (n=25,155) of the proposed cut-off (0.10) in EATV normalized to heart volumes (EATVh) compared to quartile (q1 and q4) based cut-offs for identifying obstructive CAD (defined as > 50% stenosis in any coronary artery) and high-risk individuals (defined as having a SCORE2 of ≥7.5). The positive (PPV) and negative (NPV) predictive values, the positive (+LR) and negative (-LR) likelihood ratios, sensitivity (Sens.) and specificity (Spec.) are shown. In the rightmost column, the prevalence (Prev.) for each outcome is shown. All metrics are shown with percentage (%) values.

| Sex | Outcome | EATVh | PPV (95% CI) | NPV (95% CI) | (+)LR (95% CI) | (-)LR (95% CI) | Sens. | Spec. | Prev. |
| --- | --- | --- | --- | --- | --- | --- | --- | --- | --- |
| Men | Stenosis > 50% | 0.10 | 8.8 (8.3-9.4) | 95.0 (93.3-96.4) | 1.03 (1.02-1.05) | 0.56 (0.42-0.76) | 96 | 7.2 | 8.6 |
| Women | Stenosis > 50% | 0.10 | 2.4 (2.2-2.7) | 99.2 (98.4-99.7) | 1.05 (1.03-1.07) | 0.34 (0.16-0.71) | 97.6 | 6.9 | 2.3 |
| Men | SCORE2 $\geq 7.5$ | 0.10 | 29.1 (28.3-30.0) | 90.6 (88.4-92.4) | 1.07 (1.06-1.08) | 0.27 (0.22-0.34) | 97.6 | 8.7 | 27.8 |
| Women | SCORE2 $\geq 7.5$ | 0.10 | 4.3 (3.9-4.6) | 99.3 (98.5-99.7) | 1.06 (1.05-1.07) | 0.17 (0.07-0.37) | 98.8 | 7.1 | 4 |
| Men | Stenosis > 50% | 0.14 | 9.6 (9.0-10.2) | 94.8 (93.9-95.6) | 1.13 (1.10-1.16) | 0.59 (0.50-0.69) | 86.1 | 23.7 | 8.6 |
| Women | Stenosis > 50% | 0.14 | 2.7 (2.4-3.0) | 98.8 (98.3-99.1) | 1.15 (1.10-1.21) | 0.52 (0.38-0.70) | 87.5 | 24.2 | 2.3 |
| Men | SCORE2 $\geq 7.5$ | 0.14 | 32.1 (31.2-33.0) | 86.9 (85.6-88.1) | 1.23 (1.21-1.25) | 0.39 (0.35-0.43) | 89.2 | 27.5 | 27.8 |
| Women | SCORE2 $\geq 7.5$ | 0.14 | 4.9 (4.5-5.4) | 98.9 (98.5-99.3) | 1.24 (1.21-1.27) | 0.26 (0.19-0.37) | 93.6 | 24.6 | 4 |
| Men | Stenosis > 50% | 0.25 | 12.2 (11.1-13.5) | 92.6 (92.0-93.1) | 1.49 (1.36-1.63) | 0.85 (0.82-0.89) | 34.1 | 77 | 8.6 |
| Women | Stenosis > 50% | 0.25 | 3.7 (3.0-4.4) | 98.1 (97.8-98.3) | 1.60 (1.37-1.87) | 0.83 (0.76-0.90) | 35.7 | 77.7 | 2.3 |
| Men | SCORE2 $\geq 7.5$ | 0.25 | 41.7 (39.9-43.5) | 76.6 (75.8-77.5) | 1.86 (1.75-1.98) | 0.79 (0.77-0.82) | 36 | 80.7 | 27.8 |
| Women | SCORE2 $\geq 7.5$ | 0.25 | 9.1 (8.0-10.2) | 97.5 (97.1-97.8) | 2.38 (2.17-2.61) | 0.62 (0.57-0.68) | 51 | 78.6 | 4 |

**Supplement: Table S.5.** Diagnostic performance in the clinical cohort (n=2,482) of the proposed cut-off (0.10) in EATV normalized to heart volumes (EATVh) compared to quartile (q1 and q4) based cut-offs for identifying obstructive CAD (defined as > 50% stenosis in any coronary artery). The positive (PPV) and negative (NPV) predictive values, the positive (+LR) and negative (-LR) likelihood ratios, sensitivity (Sens.) and specificity (Spec.) are shown. In the rightmost column, the prevalence (Prev.) for each outcome is shown. All metrics are shown with percentage (%) values.

| Group | Outcome | EATVh | PPV (95% CI) | NPV (95% CI) | (+)LR (95% CI) | (-)LR (95% CI) | Sens. | Spec. | Prev. |
| --- | --- | --- | --- | --- | --- | --- | --- | --- | --- |
| Men | Stenosis > 50% | 0.10 | 35.0 (32.1-38.1) | 82.6 (61.2-95.0) | 1.02 (1.00-1.03) | 0.40 (0.14-1.16) | 98.9 | 2.8 | 34.6 |
| Women | Stenosis > 50% | 0.10 | 16.1 (14.3-18.2) | 93.5 (78.6-99.2) | 1.02 (1.00-1.03) | 0.36 (0.09-1.51) | 99.1 | 2.4 | 15.9 |
| Men | Stenosis > 50% | 0.14 | 36.4 (33.4-39.6) | 83.2 (74.1-90.1) | 1.08 (1.04-1.12) | 0.38 (0.23-0.64) | 95.6 | 11.6 | 34.6 |
| Women | Stenosis > 50% | 0.14 | 16.5 (14.5-18.6) | 89.9 (83.0-94.7) | 1.04 (1.00-1.08) | 0.59 (0.33-1.06) | 94.8 | 8.8 | 15.9 |
| Men | Stenosis > 50% | 0.25 | 38.9 (34.3-43.6) | 68.5 (64.6-72.2) | 1.20 (1.04-1.39) | 0.87 (0.77-0.97) | 47.8 | 60.2 | 34.6 |
| Women | Stenosis > 50% | 0.25 | 17.6 (14.8-20.6) | 85.6 (82.9-88.0) | 1.13 (0.98-1.29) | 0.89 (0.77-1.03) | 53 | 52.9 | 15.9 |

**Supplement: Figure S.1.**
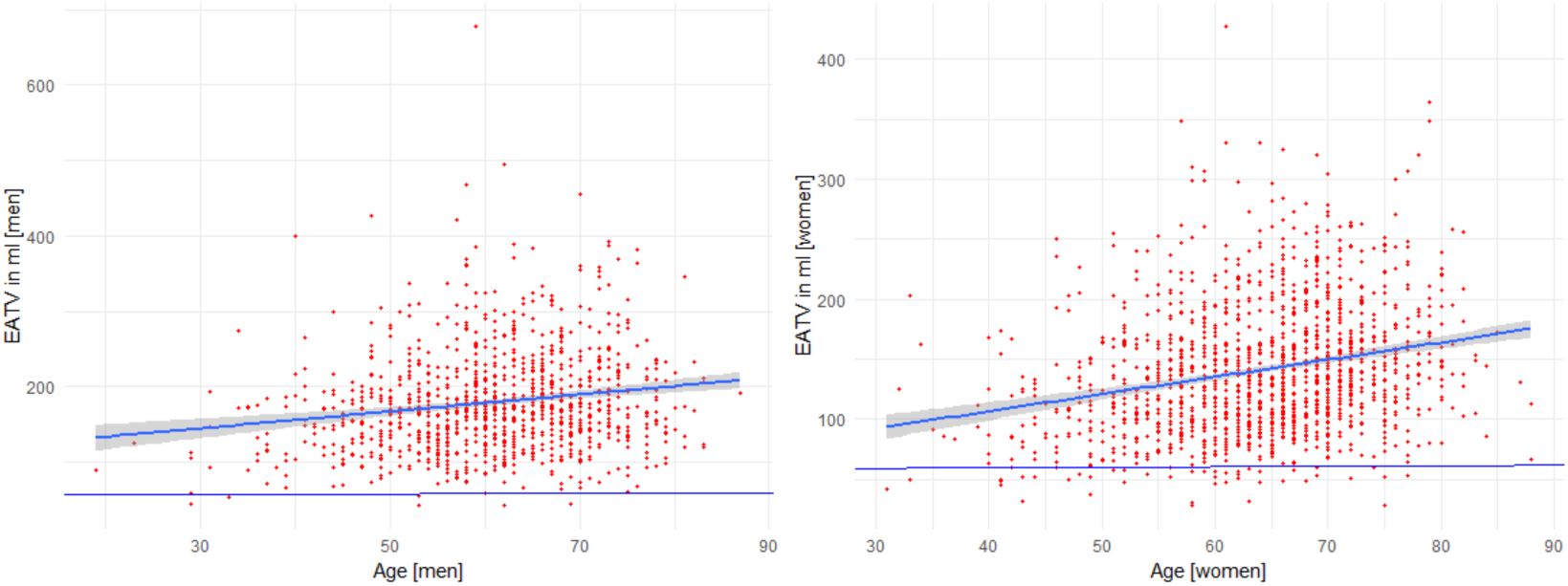
Relationship between EATV and age in men (n=1,039) and women (n=1,443) in the clinical cohort. The upper row shows non-normalized EATV and the lower row shows EATV normalized to total heart volume.

**Supplement: Figure S.2.**
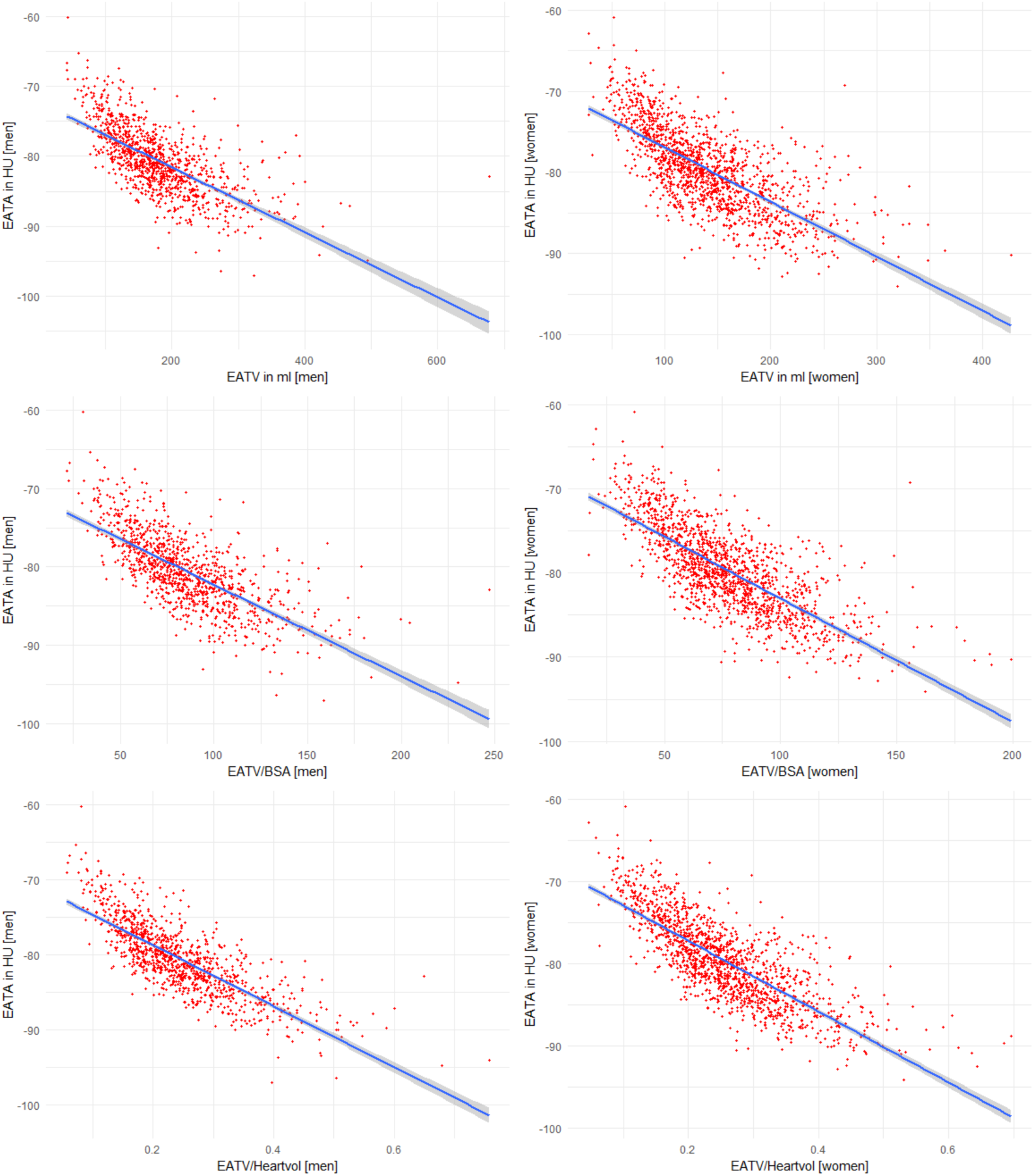
Epicardial adipose tissue attenuation (EATA) as a function of epicardial adipose tissue volume (EATV) in men (n=1,039) and women (n=1,443) in the clinical cohort. The upper row shows non-normalized EATV, the mid row EATV normalized to body surface area, and the lower row EATV normalized to heart volume.

**Supplement: Figure S.3.**
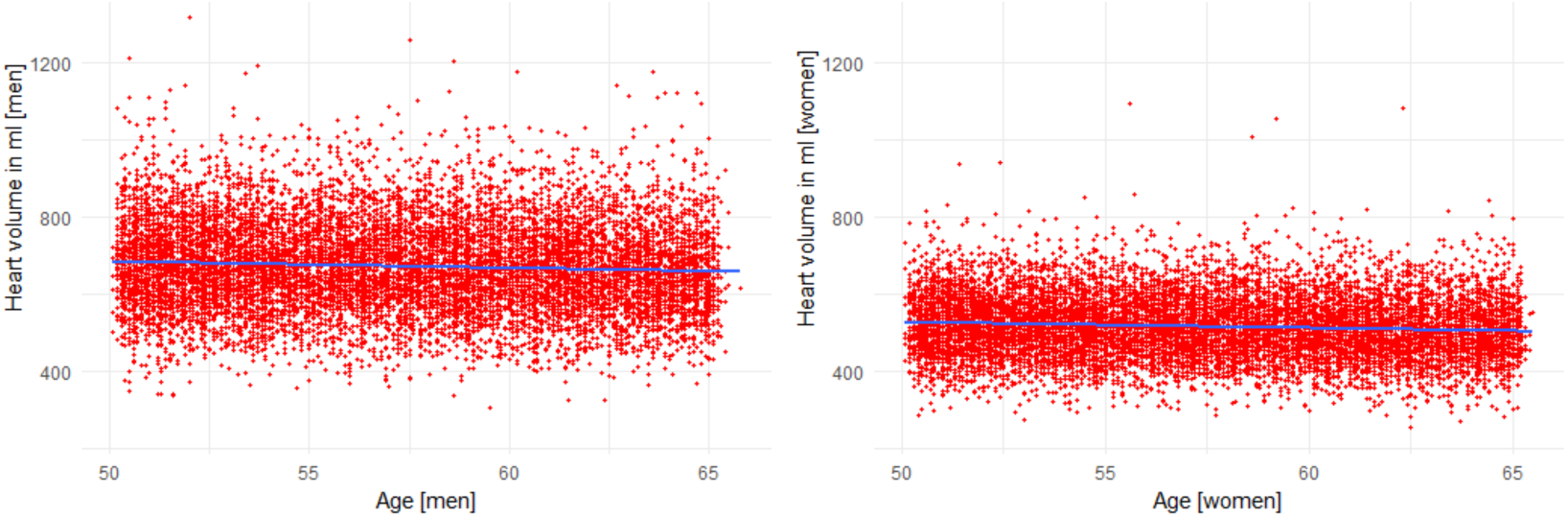
Heart volume in men (n=12,409) and women (n=12,746) in relation to age in the population cohort.

**Supplement: Figure S.4.**
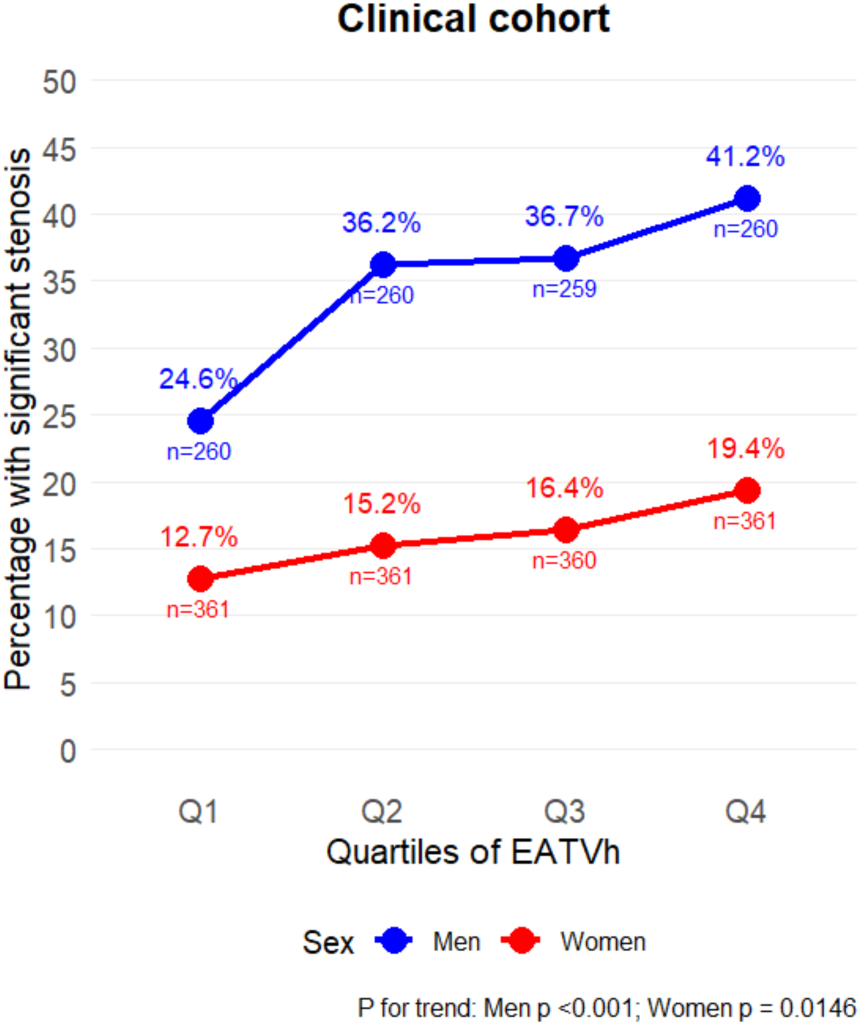
The prevalence of obstructive coronary artery disease (defined as stenosis of > 50 % in any coronary artery) by quartiles of epicardial adipose tissue normalized to heart volume (EATVh) in the clinical cohort (n=2,482). P-values for the trend over quartiles is shown beneath the plot.

